# Efficient molecular mendelian randomization screens with LaScaMolMR.jl

**DOI:** 10.1101/2024.08.29.24312805

**Authors:** Samuel Mathieu, Louis-Hippolyte Minvielle Moncla, Mewen Briend, Valentine Duclos, Anne Rufiange, Yohan Bossé, Patrick Mathieu

**Affiliations:** Quebec Heart and Lung institute, Laval University, Quebec, Canada; Department of Molecular Medicine, Laval University, Quebec, Canada; Department of Surgery, Laval University, Quebec, Canada

**Keywords:** Genetics, Mendelian Randomization, Meta-analysis, Julia Language

## Abstract

**Summary:** The ever-growing genetic cohorts lead to an increase in scale of molecular Quantitative Trait Loci (QTL) studies, creating opportunities for more extensive two samples Mendelian randomization (MR) investigations aiming to identify causal relationships between molecular traits and diseases. This increase led to the identification of multiple causal candidates and potential drug targets over time. However, the increase in scale of such studies and higher dimension multi-omic data come with computational challenges. We present “LArge SCAle MOLecular Mendelian Randomization with Julia” (LaScaMolMR.jl), an open-sourced integrated Julia package optimized for Omic-wide Mendelian Randomization (OWMR) Studies. This versatile package eliminates the two-language problem and implements fast algorithms for instrumental variable selection approaches with both cis and trans instruments and performs the most popular regression estimators for MR studies with molecular exposures. It reduces the compute time via meta-programming allowing easy deployment of multi-threaded approach and the internalization of linkage disequilibrium investigation of potential instrumental variables. Via its integrated approach and high-computational performance, LaScaMolMR.jl allows users who have minimal programming experience to perform large scale OWMR studies.

**Implementation and availability:** LaScaMolMR is freely available at github.com/SamuelMathieu-code/LaScaMolMR.jl.

## Introduction

This paper introduces a high throughput Mendelian Randomization (MR) pipeline to identify molecular risk or protective factors of partially heritable diseases. **Figure 1A** describes this pipeline and its implementation. MR is a meta-regression analysis used to infer a causal link between two phenotypes by observing the effect of single nucleotide variants (SNVs) associated with the exposure on the outcome. This technique leverages the random attribution of genetic variants to use those as instrumental variables (IVs). The selected instruments must fulfil three conditions for validity^1^ (**Figure 1B**).

**Figure 1.**
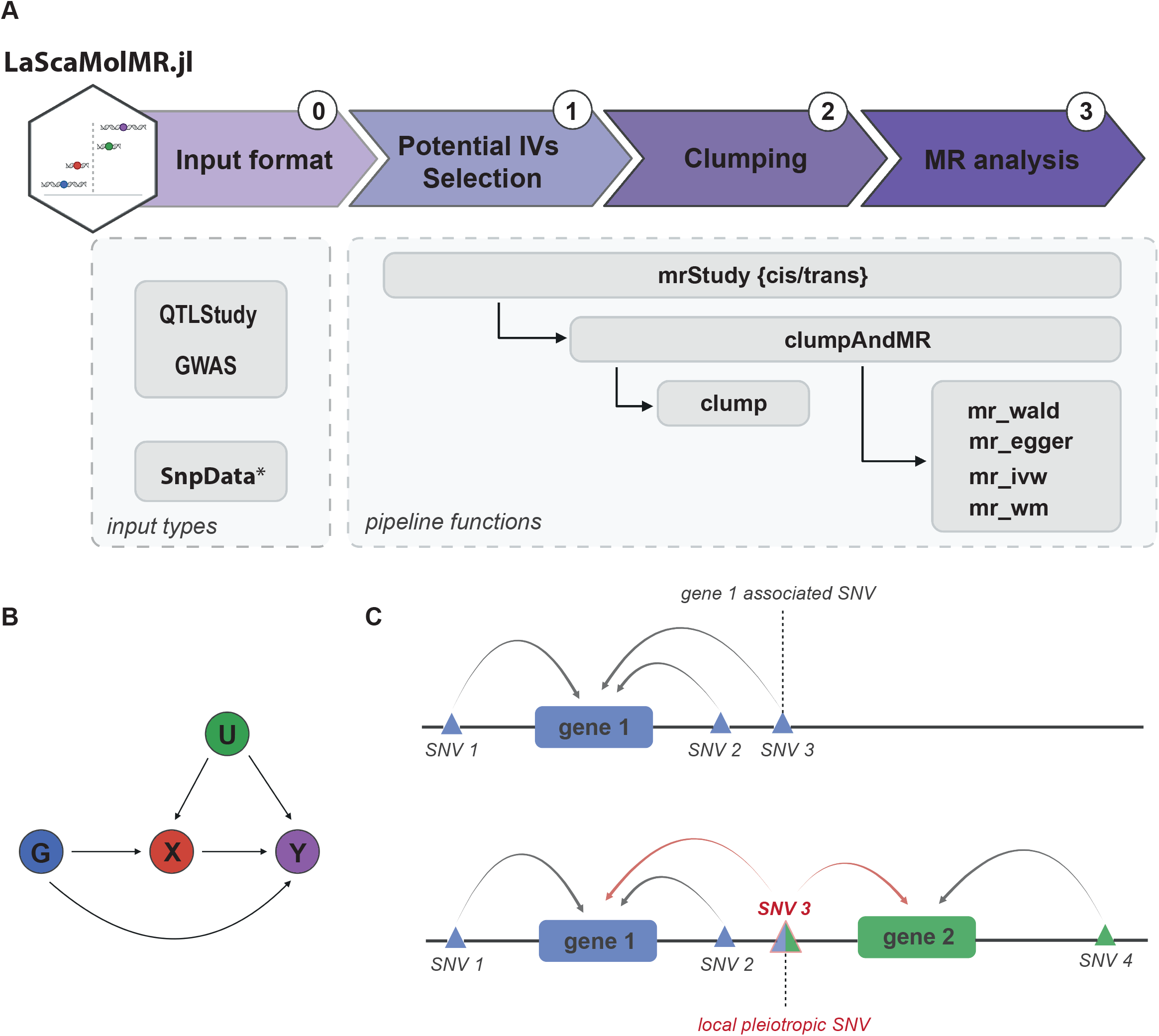
**A)** Process chart of the implemented pipeline and call diagram of the implemented and exported functions. Functions overlapping a step of the process are responsible of that step. * SnpData is provided by the SnpArrays.jl package. **B)** Directed acyclic graph schematizing MR assumptions. G is the instrument variable variant, X is the exposure phenotype, Y is the outcome phenotype and U is a confounding factor. **C)** Locus showing local pleiotropy in transcriptome-wide MR. In such setting, SNV3 would be discarded in the MiLoP approach. A multivariable MR estimator can account for measured pleiotropic effects by including SNVs 1 to 4.

1. The instrument must be associated with exposure.
2. The instrument must not be associated with a confounding factor between the exposure and the outcome.
3. The instrument must not be associated to the outcome through another variable than the exposure.

The last assumption is also known as the exclusion restriction principle. Horizontal pleiotropy consists of the violation of assumption 3. With the augmented scale of recent Genome Wide Association Studies (GWAS) and Quantitative Trait Loci studies (QTL) and their greater availability, more studies investigate molecular risk factors in large scale MR screens^2–7^. Also, more complex settings for OWMR studies such as mediation analysis between different Omic levels^8,9^ are proposed. For these reasons, robust IV selection protocols and integrated tools for such analyses are more than ever relevant. Although multivariate approaches to OWMR better account for local pleiotropy (i.e. local to effects measured in the exposure dataset)^10^, a significant proportion of studies prefer a univariate approach^2–7^ and investigate pleiotropic effects with a follow up multivariate analysis among related traits (for example genes in a locus or biologically similar traits). Although this does not allow to systematically account for pleiotropic effects within investigated exposures, it simplifies the pipeline and benefits from the statistical power of the univariate setting. We thus implement a univariate solution to the problem while allowing the user to remove IVs potentially associated to multiple targets. This approach limits the study to direct and genetically independent effects. Significant associations among pleiotropic loci or groups of traits are encouraged to be further investigated using a multivariate MR estimator^11,12^ or a functional analysis.

Julia is a free open-sourced scientific computing programming language. It uses a Just in Time (JIT) compiler that compiles a function at first call. This allows powerful meta-programming macros modifying expressions before compilation. Julia packages thus often show outstanding performance while offering a high-level interface to users. This allows to address what is known as the two-language problem or the tendency of scientific code to use a slow but flexible language for frontend (such as R or Python) and a fast but less flexible language in backend (such as Rust or C). This widespread practice makes development less accessible to scientists and adds complexity to code optimization process.

## Methods

### The Mendelian Randomization pipeline

LaScaMolMR provides two data types describing the structure of genetic association tabular data (’GWAS’ and ‘QTLStudy’) providing a high-level interface to file parsing. ‘QTLStudy’ describes association data with multiple targets. Files can be stored in different folders or files depending on chromosome or target name, thus allowing multiple formats depending on the usage (Study among selected targets/all targets, cis/trans instruments). Handling of tabular data is done using the InMemoryDatasets.jl and DLMReader.jl Julia packages. These threaded packages implement fast file parsing and data manipulation functions.

The pipeline is divided in three steps (**Figure 1A**). In the first step, files are parsed according to structure defined in ‘GWAS’ and ‘QTLStudy’ objects in the function ‘mrStudy’. ‘mrStudy’ also takes a ‘type’ argument which corresponds to cis or trans IV selection. Cis potential IV filtering includes 3 filters:

1. Filter out variants for which the association p-value to the exposure is higher than ‘p_tresh’
2. Filter out variants further than window base pairs from the exposure trait (typically gene transcription starts site)
3. Filter out indels and non-biallelic variants.

Trans selection only includes the first and the third filter. Other filters can be applied by the user to filter out rare variants, instruments which might induce reverse causality, or local pleiotropic variants. The “Mitigated Local Pleiotropy” (MiLoP) approach filters out potential IVs which are associated to a second target at ‘p_tresh_MiLoP’ level (**Figure 1C**), in opposition to the said “naive” approach which does not include a local pleiotropy assessment. Both approaches are implemented in the package although “naive” is the default.

In the second section of the pipeline, independent IVs are selected among potential IVs (**Figure 1A**). In order to assess linkage disequilibrium (LD) between potential IVs, composite LD^13^ is calculated and fed into a clumping algorithm (**Algorithm 1**). The user must provide a reference panel in Plink 1.9 format. The reference data is parsed with SnpArrays.jl package brought by the OpenMendel project^14^. The function ‘ClumpAndMR’ takes a dataset of potential IVs grouped by exposure and performs clumping and MR analysis for each exposure. This function can also provide detailed information relative to chosen IVs. The documentation provides detailed information to that matter.

The last section of the pipeline is the MR analysis. The package implements 4 popular MR estimators. These include Inverse Variance Weighted, Weighted Median, Egger and the Wald ratio. While the 3 first methods aim at assessing a causal relationship, the Wald ratio (or any single IV approach) cannot distinguish causality from pleiotropy^15^. All MR implementations share a common interface which facilitates customization of the pipeline through user defined functions. Each function takes 4 vectors of entry and have a ‘α’ option corresponding to the α-value of the confidence intervals. The ‘mr_output’ type structures the outputs with a common interface. This output includes summary data of association as well as sensitivity tests such as the Cochran’s Q test for heterogeneity and the Egger intercept. These tests are commonly used to control non-measured horizontal pleiotropic effects.

### Multi-threading

The multi-threading in the pipeline section 1 is provided entirely by parsing, filtering and grouping functions of InMemoryDatasets and DLMReader. In the section 2, the clumping function is threaded using the ‘@threads’ macro (**Algorithm 1**) Julia macros offer a high-level interface to multi-threading with similar capabilities and performance to the OpenMP compilation instructions in C/C++^16^.

## Results

### Large scale benchmark

We ran the full pipeline using Coronary Artery Disease (CAD) GWAS^17^ (GCST90132314) as outcome and GTEx V8 eQTL data from all 49 tissues as exposure^18^. Files were not previously filtered. Analysis for each tissue took an average time of 20 minutes and 13 seconds with 10 threads on an Ubuntu 20.04 server with 160 Intel(R) Xeon(R) Gold 6230 CPU @ 2.10GHz processors and 754G of DDR4 ram at 2933 MT/s **(Supplementary Table 1)**. A total of 238528 (gene, tissue) pairs were investigated, 22417 of which had at least 3 IVs (**Supplementary Table 2**). The analysis identified 2342 candidate (gene, tissue) pairs after FDR 5% adjustment and further removing targets which had non-zero Egger intercept or showed heterogeneity (**Supplementary Table 3)**. These associations target 811 different genes, thus highlighting the polygenic character of the disease. We also ran a simplified analysis using only Coronary Artery tissue cis-eQTLs using 10 threads on an Ubuntu 22.04 desktop with 16 Intel(R) Core(TM) i7-10700K CPU at 3.80GHz processors and 84GB of DDR4 RAM at 3200MHz. This analysis took about 14 minutes and peaked at around 45 GB of memory usage. Of note: using the ‘mrStudyNFolds’ function can help reduce the memory load.

### Comparing Naive results to MiLoP results

We performed a Proteome-wide MR study using blood protein levels from the Fenland study^19^ (https://www.synapse.org/Synapse:syn51761394/wiki/622766) as exposures and CAD^17^ as outcome. Among many hits with the naive approach, some proteins belong to genes in the same locus, such as ITIH1/ITIH3 and ERAP1/ERAP2. These effects are driven by locally pleiotropic variants. Using the MiLoP approach allows to restrict the findings to genetically independent associations. In both locus, variants associated to both proteins at p<0.05 level were discarded. Nominal significance was lost for all proteins except ITIH3 (**Supplementary Table 4, Supplementary Figure 1**).

### Clumping function benchmark

As the complexity of the clumping algorithm is quadratic with respect to the number of variants and linear with the number of individuals in the reference panel (*O*(*m*^2^*n*) in worse case), the scaling of this component of the pipeline is a challenge. We built a lightweight implementation by using powerful Julia macros such as ‘@simd’ and ‘@unbound’ backed with multi-threading. Most pipelines use Plink 1.9^20^ for variant clumping^5,6^. This solution does not scale well to OWMR as reference genotype files are parsed at every query. It also involves multiple file writings and readings. Our implementation replicates Plink results while avoiding this burden. We benchmarked the function using real data from Fenland. Clumping 2797 variants with Plink using 10 threads took 2.27 seconds (not accounting for the time to read clumped file) while LaScaMolMR’s clump took 1.05 seconds when considering bimbedfam file parsing and 0.02 seconds otherwise. In practice, compute time is found to scale linearly with the number of variants. Also, optimal performance is achieved with 10 to 16 processor cores **(Supplementary Table 5, Supplementary Figure 2)**. Efficient parallelization of this algorithm when more resources are available remains a challenge.

## Conclusion

In short, LaScaMolMR.jl takes care of every part of a Mendelian Randomization molecular screen pipeline. The threaded Julia implementation and the internalization of linkage disequilibrium calculations enable a minimal compute time while addressing the two-language problem and offering the user a high-level flexible interface. Such a tool can serve as a baseline in the future for more complex study settings such as those involving mediation analysis or a better local horizontal pleiotropy assessment.

## Data Availability

All data used is publicly available at provided urls in section "Declarations". Code and documentation links are at "Data Availability links" section.

https://github.com/SamuelMathieu-code/LaScaMolMR.jl

https://samuelmathieu-code.github.io/LaScaMolMR/

## Author contributions statement

S.M., L.H.M.M., M.B. and P.M. elaborated the pipeline. S.M. wrote the software. S.M. and P.M. drafted the manuscript. L.H.M.M. and S.M. drafted the figures. S.M., L.H.M.M., M.B., V.D., A.R., Y.B. and P.M. revised the manuscript and provided constructive critics.

## Acknowledgments

Work of the authors is supported by the Canadian Institutes of Health Research grants to P.M. (FRN159697, FRN191807) and the Quebec Heart and Lung Institute Fund. Y.B. holds a Canada Research Chair in Genomics of Heart and Lung Diseases P.M. is the recipient of the Joseph C. Edwards Foundation granted to Université Laval. S.M. and M.B. are supported by the Fonds de Recherche du Québec secteur Santé through a scholarship.

## Software and data availability

All data used for the example case is public and freely available. Code for LaScaMolMR and all its dependencies is open-source. Code from LaScaMolMR is available at github.com/SamuelMathieu-code/LaScaMolMR.jl. Documentation is available at samuelmathieu-code.github.io/LaScaMolMR/.

