## Supplementary Figure 1 for "Efficient molecular mendelian randomization screens with LaScaMolMR.jl"

### Supplementary Figure 01

| Blood proteins | IVs | Odds ratio | OR (95% CI) IVW | Log P-values | P-values IVW |
|---|---|---|---|---|---|
| **Locus ERAP** | | | | | |
| ERAP1.4964.67 | 10 | | 0.99 (0.98 to 0.99) | | 2.35e-04 |
| (MiLoP) ERAP1.4964.67 | 5 | | 0.99 (0.98 to 1.00) | | 1.77e-01 |
| ERAP2.8960.3 | 16 | | 1.02 (1.01 to 1.02) | | 4.50e-05 |
| (MiLoP) ERAP2.8960.3 | 5 | | 1.00 (0.97 to 1.03) | | 9.73e-01 |
| **Locus ITIH** | | | | | |
| ITIH1.7955.195 | 10 | | 0.97 (0.96 to 0.99) | | 2.50e-05 |
| (MiLoP) ITIH1.7955.195 | 2 | | 1.01 (0.97 to 1.06) | | 5.35e-01 |
| ITIH3.7145.1 | 9 | | 1.04 (1.03 to 1.06) | | 5.01e-07 |
| (MiLoP) ITIH3.7145.1 | 2 | | 1.07 (1.02 to 1.11) | | 1.78e-03 |

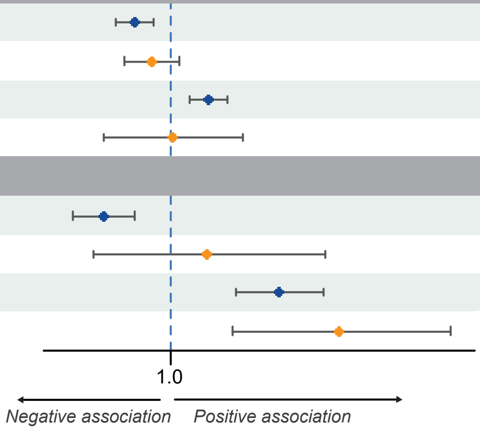


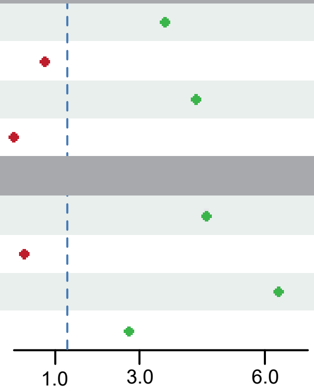
