## Supplementary figures and images for "Efficient molecular mendelian randomization screens with LaScaMolMR.jl"

### Supplementary Figure 2

**Supplementary Figure 02**

**A**

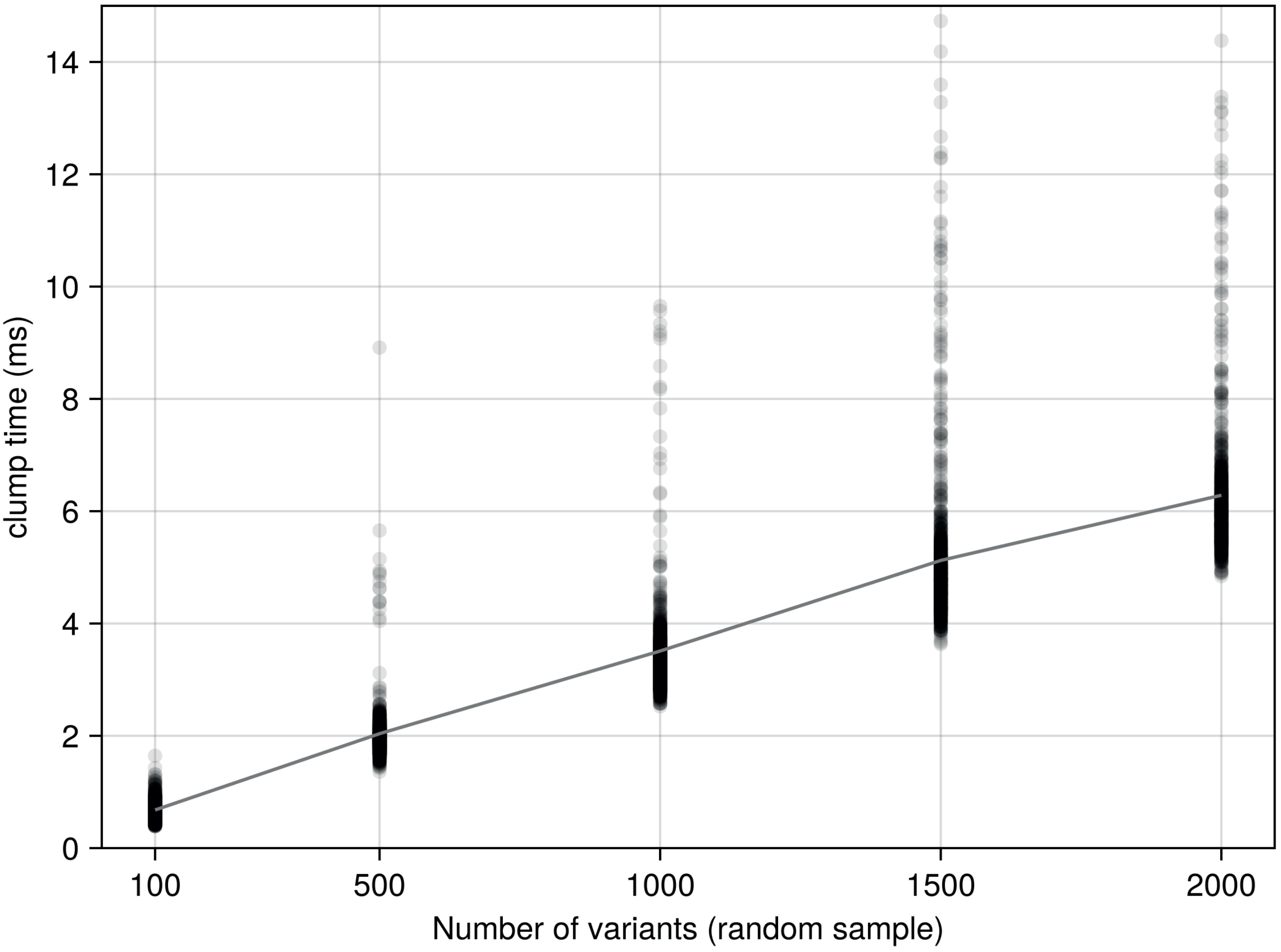


**B**

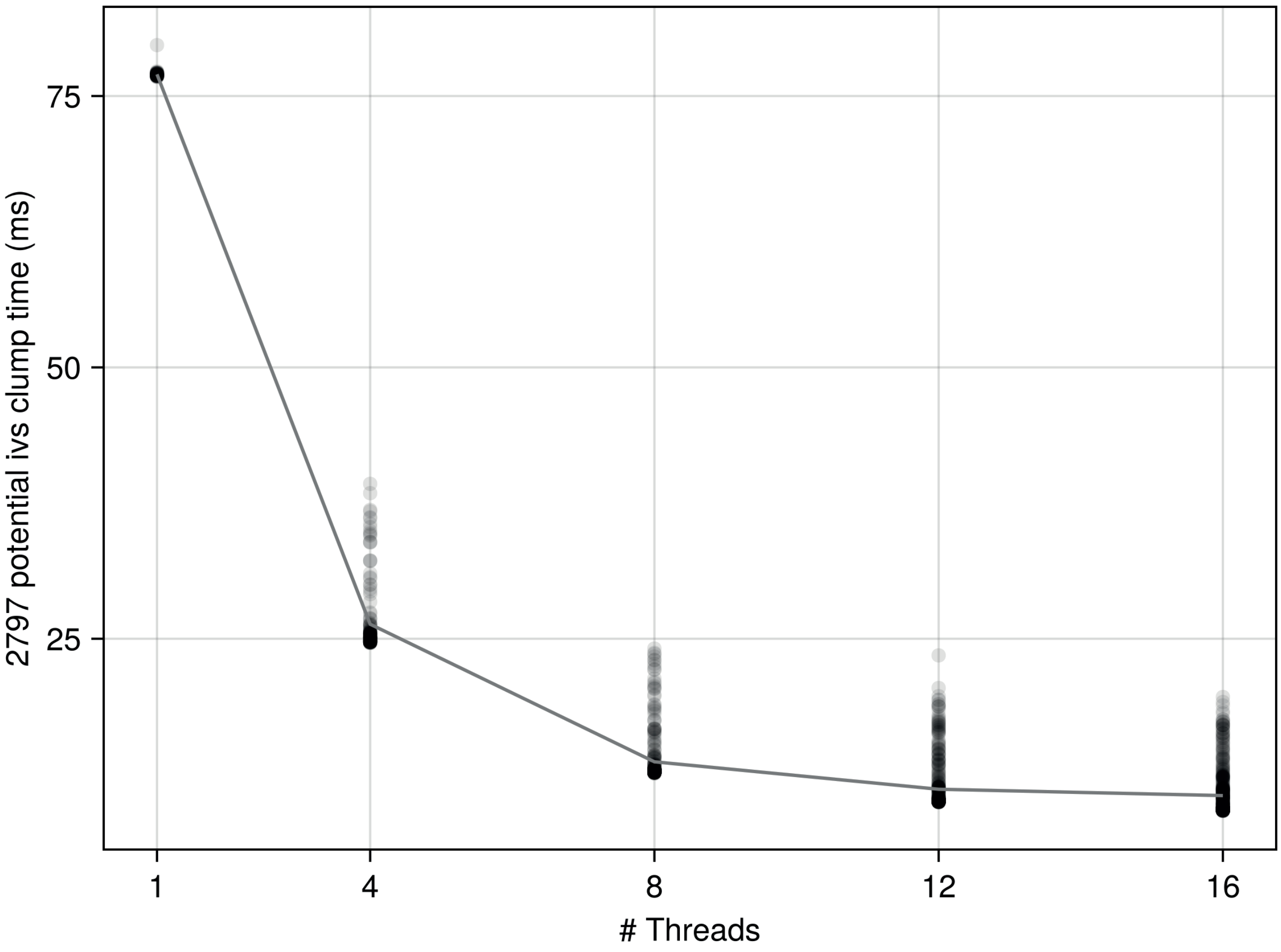
